# A novel missense variant in the ATPase domain of ATP8A2 and review of phenotypic variability of *ATP8A2*-related disorders caused by missense changes

**DOI:** 10.1101/2024.05.15.24306843

**Authors:** Kyle P. Flannery, Sylvia Safwat, Eli Matsell, Namarata Battula, Ahlam A. A. Hamed, Inaam N. Mohamed, Maha A. Elseed, Mahmoud Koko, Rayan Abubaker, Fatima Abozar, Liena E. O. Elsayed, Vikram Bhise, Robert S. Molday, Mustafa A. Salih, Ashraf Yahia, M. Chiara Manzini

## Abstract

ATPase, class 1, type 8A, member 2 (ATP8A2) is a P4-ATPase with a critical role in phospholipid translocation across the plasma membrane. Pathogenic variants in *ATP8A2* are known to cause cerebellar ataxia, mental retardation, and disequilibrium syndrome 4 (CAMRQ4) which is often associated with encephalopathy, global developmental delay, and severe motor deficits. Here, we present a family with two siblings presenting with global developmental delay, intellectual disability, spasticity, ataxia, nystagmus, and thin corpus callosum. Whole exome sequencing revealed a homozygous missense variant in the nucleotide binding domain of ATP8A2 (p.Leu538Pro) that results in near complete loss of protein expression. This is in line with other missense variants in the same domain leading to protein misfolding and loss of ATPase function. In addition, by performing diffusion-weighted imaging, we identified bilateral hyperintensities in the posterior limbs of the internal capsule suggesting possible microstructural changes in axon tracts that had not been appreciated before and could contribute to the sensorimotor deficits in these individuals.

## Introduction

Biallelic mutations in *ATPase, class 1, type 8A, member 2* (*ATP8A2*) (MIM:605870) cause <u>c</u>erebellar <u>a</u>taxia, <u>m</u>ental retardation, and dise<u>q</u>uilibrium syndrome <u>4</u> (CAMRQ4). The classic presentation of CAMRQ4 is characterized by intellectual disability, ataxia, dysarthria, and occasionally quadrupedal gait (Alsahli et al., 2018). However, the spectrum of phenotypes caused by ATP8A2 loss of function is variable due to the broad expression pattern of this gene in the brain and eyes.

*ATP8A2* encodes a P4-ATPase that forms a complex with CDC50A to translocate phospholipids phosphatidylserine (PS) and phosphatidylethanolamine (PE) from the outer (exoplasmic) layer of the membrane to the inner cytoplasmic layer, thus maintaining asymmetry in the composition of the lipid bilayer (Coleman & Molday, 2011; Tadini-Buoninsegni et al., 2019; Vestergaard et al., 2014). Phospholipid asymmetry confers different structural and functional properties to the outer and inner sides of the plasma membrane (Lorent et al., 2020). For example, PS in the exoplasmic layer of the cell membrane triggers phagocytosis of apoptotic cells controlling cell survival (Fadok et al., 1992; Martin et al., 1996). In addition to apoptosis, exoplasmic PS is also known to play a vital role in physiological processes including blood coagulation (Bevers et al., 1982) and myotube formation (van den Eijnde et al., 2001).

ATP8A2 is expressed in many brain regions, with highest expression observed in the cerebellum (Onat et al., 2013) and the hippocampus (Cacciagli et al., 2010). The expression pattern strongly correlates with structural brain malformations observed in patients with pathogenic *ATP8A2* variants including atrophy of the cerebral cortex, corpus callosum, and cerebellum (Martín-Hernández et al., 2016; Onat et al., 2013). However, not all patients with pathogenic *ATP8A2* variants present with overt brain abnormalities (Damásio et al., 2021). In fact, one study reported normal magnetic resonance imaging (MRI) of the brain in 50% of patients (McMillan et al., 2018), while another noted normal brain MRI in 3/3 patients (Alsahli et al., 2018). ATP8A2 is also strongly expressed in the retina (Cacciagli et al., 2010; Tadini-Buoninsegni et al., 2019), and some patients have presented with impaired visual acuity, nystagmus, ophthalmoplegia, and bilateral optic disk atrophy (Alsahli et al., 2018; Martín-Hernández et al., 2016).

Rodent models of *Atp8a2* loss of function have recapitulated many aspects of CAMRQ4 that are observed in humans and uncovered underlying processes that may contribute to disease phenotypes. Wabbler-lethal (*wl*) mutant mice, which harbor mutations in *Atp8a2*, exhibit reduced survival, reduced body weight, abnormal gait characterized by dragging of the hind feet, hind limb clasping indicative of an underlying neurological deficit, and distal axonal degeneration (Zhu et al., 2012), the latter of which corroborated an earlier electron microscopy study of the *wl* mouse nervous system (Luse et al., 1967). Further evidence for the role of ATP8A2 in axons was uncovered in rats where overexpression of ATP8A2 was found to increase axon length in hippocampal neurons (Xu et al., 2012). The visual system was also strongly affected in *wl* mice where photoreceptor proteins were found to be correctly localized, but a reduction in the number of photoreceptors in the retina as well as the length of the outer segment layer was noted (Coleman et al., 2014). Degeneration of the optic nerve was also observed in *wl* mice (Carroll et al., 1992).

Here, we present the case of two siblings with a novel homozygous missense variant in *ATP8A2* identified through whole exome sequencing (WES). Both siblings exhibited classic CAMRQ4 symptoms including dysarthria, spasticity, and delayed motor milestones. Additionally, novel brain MRI findings of bilateral hyperintensities of the posterior limbs of the internal capsule, as well as thinned corpora callosa were observed in each case. As such, this case study further expands our knowledge of the phenotypic spectrum caused by biallelic mutations in *ATP8A2*.

## Methods

### Subjects

Ethics approval was received from the Ethical Committee of Medical Campus, University of Khartoum, Sudan, the Ethical Committee of the National University, Sudan (approval number NU-RECG200), and the Institutional Review Board at Rutgers University. Each patient and their parents were fully informed of the purpose and procedures of this study and written consent was obtained before participation. Detailed personal data, age of onset, pregnancy, and delivery history as well as family history of similar condition or any other genetic disorders with 3-generation pedigree construction were completed. Physical examination, full neurological assessment and anthropometric measurements were performed in the initial and follow-up visits. Ophthalmological examination and MRI of the brain were also performed.

### Whole exome sequencing and variant analysis

WES was performed on DNA obtained from the siblings and their unaffected parents at the Broad Institute Genomic Services (Broad Institute, Cambridge, MA). Reads were aligned to hg38 with BowTie2 (Langmead & Salzberg, 2012), and variants were called with SAMtools (Li et al., 2009) and GATK (Van der Auwera et al., 2013). Variant call format (VCF) files were then annotated with ANNOVAR (Wang et al., 2010) and stored in a custom SQL database. Homozygous variants likely to affect gene function (nonsense, missense, frameshift deletion/insertion, non-frameshift deletion/insertion, start loss, stop loss) were filtered for allele frequencies less than 1% in relevant populations in the Genome Aggregation Database (Karczewski et al., 2020) and the Greater Middle Eastern Variome (Scott et al., 2016). SIFT (Ng & Henikoff, 2003), PolyPhen2 (Adzhubei et al., 2013), CADD (Rentzsch et al., 2019), and REVEL (Pejaver et al., 2022) were used to assess the pathogenicity of missense variants. Shared inheritance and heterozygosity in each parent were confirmed in the variant filtering process.

### Plasmid construct design

The human ATP8A2 construct was previously cloned into a pcDNA3.1 plasmid engineered to contain a C-terminal 1D4 tag (Choi et al., 2019; Guissart et al., 2020). The p.Leu538Pro mutant construct was developed using site directed mutagenesis with overlapping primers designed to incorporate the missense mutation into the wildtype (WT) ATP8A2 construct using Phusion polymerase. Mutations were verified by Sanger sequencing and re-ligated into the WT plasmid using the KPN1 and Xba1 restriction enzymes.

### Cell culture and protein expression

HEK293T cells were cultured in DMEM (Sigma-Aldrich) supplemented with 8% bovine growth serum (Thermo Scientific), 100 IU/ml penicillin, 100 μg/ml streptomycin, and 2 mM l-glutamine. The cells grown on 10 cm plates to ∼50% confluency were co-transfected with 5 μg each of the ATP8A2 plasmid and CDC50A plasmids and 30 μg of polyethylenimine (PEI) per plate. The cells were harvested after 24h and pelleted at 2000 rcf (Sorval LegendRT). The cells were resuspended in lysis buffer (150mM NaCl, 50mM HEPES/NaOH pH 7.5, 5mM MgCl_2_, 20 mM CHAPS, 1x Protease Arrest, 1mM dithiothreitol (DTT), 0.5mg/mL DOPC) with stirring at 4°C. After 30 min, the detergent insoluble membrane fraction was removed by centrifugation at 40,000 rcf for 12 min. Protein expression was measured by western blotting analysis labeled with the Rho1D4 antibody and normalized to a tubulin loading control as previously described (Choi et al., 2019).

## Results

### Case reports

Here, we present 2 siblings born to healthy parents (**Fig.1A**). The proband (P1) presented with ataxia and severe global developmental delay. The pregnancy and delivery history were uneventful. At 0-5 years, P1 was hypotonic and areflexic, lacked head control, and was unable to crawl or speak. By 6-10 years of age, P1 had dysarthria, nystagmus, and dysmetria, with greater interest in their surroundings, and ambulation with unilateral support. Hearing and vision remained intact. Further examination revealed microcephaly, spasticity, and hyperreflexia of both upper and lower limbs, in addition to feet deformity. Brain MRI showed a mildly thinned corpus callosum and mildly dilated lateral ventricles (**Fig.1B-C**), but the cerebellum was intact. The diffusion-weighted imaging (DWI) sequences showed bilateral symmetric hyperintensities in the posterior limbs of the internal capsules (**Fig.1D**). P1’s sibling (P2) presented also with delayed milestones. At 0-5 years old, P2 had dysarthria and could walk with support. Microcephaly, nystagmus, and spasticity of upper and lower limbs were present, but there was no ataxia. MRI of the brain showed a thinned corpus callosum and bilateral symmetric DWI hyperintense signals in the posterior limbs of the internal capsules (**Fig.1E**).

**Figure 1.**
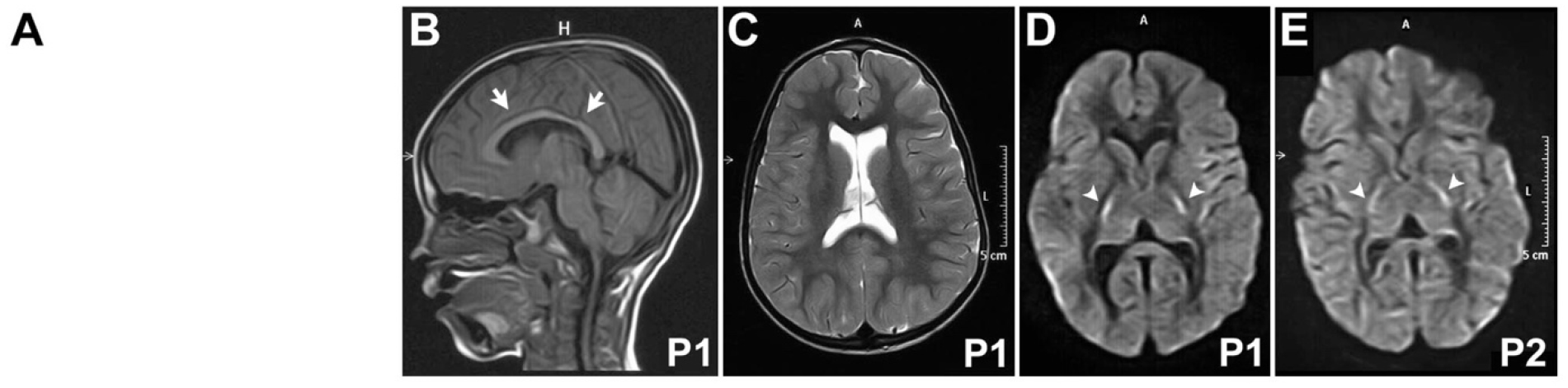
Brain imaging shows internal capsule alteration in both siblings. **A**. Family pedigree. **B-C**. Brain MRI of P1 shows thinning of the corpus callosum (arrows) in T1-weighted imaging (**B**.) and a moderate enlargement of the lateral ventricles evident in T2-weighted imaging (**C**.). **D-E**. Both siblings (P1 in **D**. and P2 in **E**.) show bilateral hyperintensity in the posterior limbs of the internal capsule (arrowheads) in diffusion-weighted imaging.

### Whole exome sequencing identified a candidate ATP8A2 variant

After WES, bioinformatic processing, and variant filtering, a total of four homozygous variants were shared between the siblings with confirmed inheritance from each parent. One missense variant was detected in *DNAH2* that was predicted by SIFT, PolyPhen2, and CADD to be damaging (SIFT: 0; PolyPhen2: 1; CADD: 27.5; REVEL: 0.775). However, biallelic mutations in *DNAH2* cause spermatogenic failure and are not known to be associated with central nervous system pathology (Li et al., 2019; Li et al., 2021). As such, this variant was ruled out as a possible candidate for the neurological presentation of the siblings. Two additional homozygous variants in the pseudogene *TPRXL*, one missense and one non-frameshift insertion, were not considered upon variant filtering.

Only a missense variant in *ATP8A2* (NM_016529.6:c.1613T>C; NP_057613.4:p.Leu538Pro), which is highly expressed throughout the brain (Cacciagli et al., 2010; Onat et al., 2013) and is a known cause of CAMRQ4, remained. This variant was strongly predicted to be deleterious by SIFT, PolyPhen2, CADD, and REVEL (SIFT: 0.001; PolyPhen2 HumDiv: 0.999; PolyPhen2 HumVar: 0.987; CADD: 28.4; REVEL: 0.781) and has never been reported in the literature or in ClinVar. This variant occurred in the cytoplasmic nucleotide binding (N) catalytic domain (**Fig. 2A**), specifically within an alpha helical secondary structure (**Fig. 2B**). Interestingly, it occurred at an amino acid residue that is highly conserved among other species but not most other P4-ATPases where amino acids with hydrophobic side chains are present (**Fig. 2C**). Using FoldX5 (Schymkowitz et al., 2005), which has been used in a previous study to predict the effect of point mutations on the overall stability of ATP8A2 disease variants (Matsell et al., 2024), we found that the proline substitution at this position likely causes a break in the helical segment leading to severe protein misfolding. Our *in vitro* studies supported this prediction, as the p.Leu538Pro variant resulted in a severe reduction in protein expression (**Fig. 2D-E**), a finding consistent with numerous other ATP8A2-disease-causing variants (Choi et al., 2019; Guissart et al., 2020; Matsell et al., 2024). Based on our findings, the American College of Medical Genomics (ACMG) guidelines for variant pathogenicity interpretation indicate that this variant is pathogenic based on one strong, two moderate, and two supporting criteria (**Table 1**) (Richards et al., 2015).

**Table 1.** ACMG criteria supporting the classification of the p.Leu538Pro variant as likely pathogenic.

| ACMG Evidence Class | Description | Justification for this Case |
| --- | --- | --- |
| PS3 (Strong) | Well established *in vitro* or *in vivo* functional studies supportive of a damaging effect on the gene or gene product. | The p.Leu538Pro variant causes nearly complete loss in ATP8A2 expression. |
| PM1 (Moderate) | Located in a mutational hotspot and/or critical and well-established functional domain (e.g. active site of an enzyme) without benign variation. | Our cases and others confirm that the catalytic cytoplasmic N and P domains are mutational hotspots. |
| PM2 (Moderate) | Absent from controls (or at extremely low frequency if recessive) in the Genome Aggregation Database. | The L538P variant is absent from the Genome Aggregation Database. |
| PP2 (Supporting) | Missense variant in a gene that has a low rate of benign missense variation and in which missense variants are a common mechanism of disease. | There are 177 missense variants in ATP8A2 reported in ClinVar, of which 3 are curated benign and 5 are likely benign. **Date Accessed: 26 April 2024** |
| PP3 (Supporting) | Multiple lines of computational evidence support a deleterious effect on the gene or gene product (conservation, evolutionary, splicing impact, etc.). | SIFT, PolyPhen, CADD, REVEL, and FoldX5 all support a deleterious effect of L538P on ATP8A2. |

**Figure 2.**
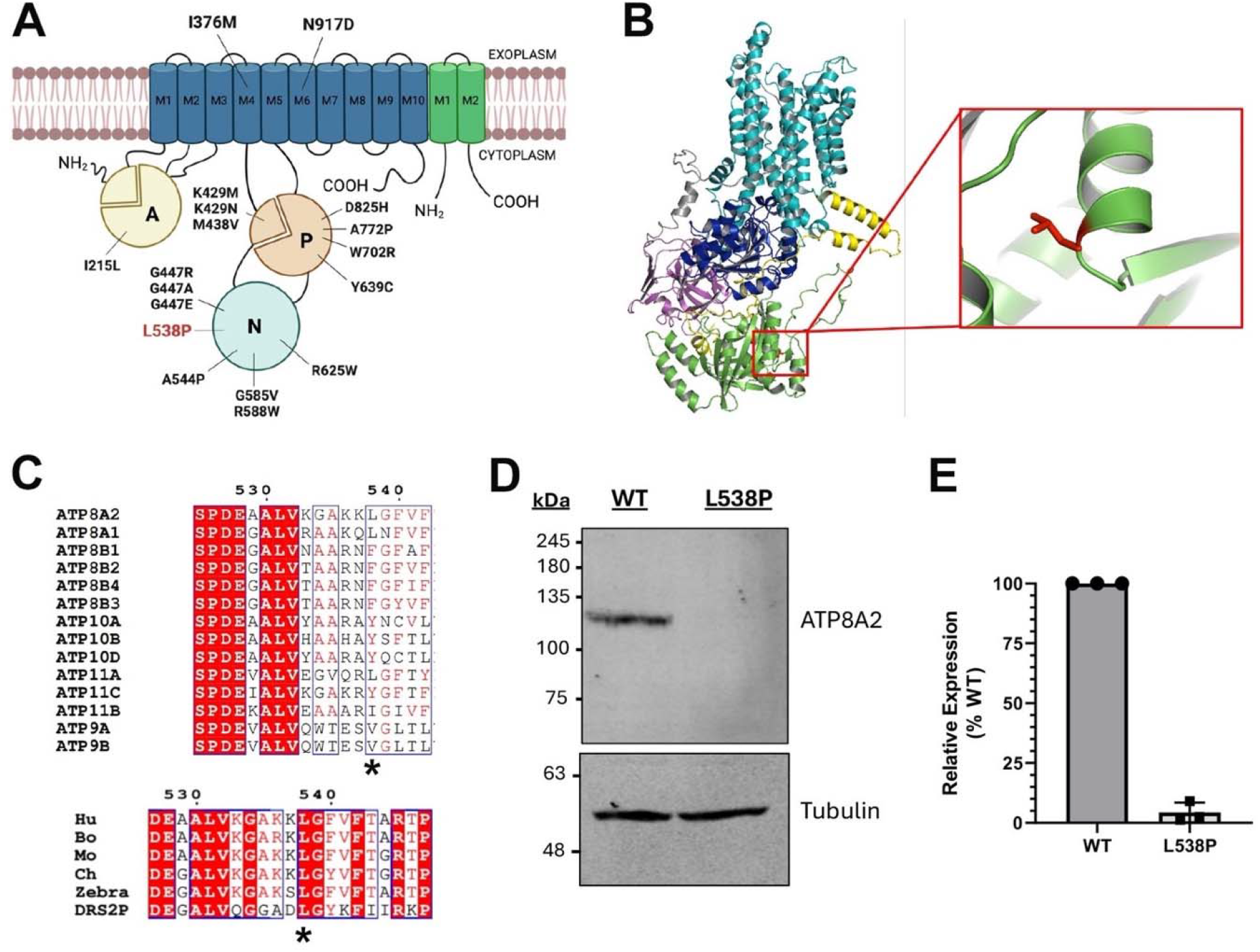
Conservation and localization of the p.Leu538Pro (L538P) variant. **A**. L538P is located in the nucleotide binding (N) domain. Most previously identified missense variants in ATP8A2 occur in the cytoplasmic N, actuator (A), or phosphorylation (P) catalytic domains. **B**. Alphafold structure of ATP8A2 shows that the L538P mutation is located within an alpha helical region of the nucleotide binding domain (N-domain). **C**. This residue is highly conserved within different species of ATP8A2 (Human (Q9NTI2) Bos taurus (C7EXK4), Mus musculus (P98200), Gallus gallus (A0A1D5P6U3), Danio rerio (A0A8M1RFW8) and the yeast homolog DRS2P (P39524) but less conserved in other P4-ATPases that all contain an amino acid with a hydrophobic side chain. **D-E**. Western blot analysis of the mutant construct reveals little if any expression of the L538P variant compared to that of WT ATP8A2 (WT: N=3; L538P: N=3).

## Discussion

*ATP8A2* mutations have been identified as the underlying cause of CAMRQ4 syndrome which is characterized by encephalopathy, intellectual disability, severe hypotonia, psychomotor delay, chorea, and optic atrophy (Alsahli et al., 2018; Cacciagli et al., 2010; Damásio et al., 2021; Guissart et al., 2020; Martín-Hernández et al., 2016; McMillan et al., 2018; Narishige et al., 2022; Onat et al., 2013). Additionally, individuals with *ATP8A2* mutations may exhibit other neurological manifestations such as tremors, seizures, and/or abnormal brain imaging, especially cerebellar atrophy (Guissart et al., 2020). The patients described in this study harbor a novel, homozygous missense variant (p.Leu538Pro) at a highly conserved amino acid in *ATP8A2* that results in protein degradation, likely due to severe misfolding. Missense variation in the cytoplasmic catalytic domains, primarily the nucleotide binding (N) and phosphorylation (P) domains, has become an increasingly common mechanism of ATP8A2-related disease (Choi et al., 2019; Guissart et al., 2020; Heidari et al., 2021; Martín-Hernández et al., 2016; Matsell et al., 2024; McMillan et al., 2018; Onat et al., 2013). As such, we classify this variant as pathogenic based on ACMG criteria.

The cytoplasmic N, P, and actuator (A) catalytic domains are well conserved in P-type ATPases, with the N domain acting as a kinase, the A domain acting as a phosphatase, and the P domain containing an aspartic acid residue (Asp428) that the N and A domains act on (Palmgren & Nissen, 2011). Upon Asp428 phosphorylation, PS and PE are flipped from the exoplasmic leaflet to the cytoplasmic leaflet of the plasma membrane (López-Marqués et al., 2020). Multiple studies have focused significant attention on missense variants in these catalytic domains and their effects on protein expression, localization, and ATPase activity. Most reported missense variants are in the N and P domains (**Fig.2B**) where they disrupt ATPase activity (P: p.Lys429Met, p.Met438Val; N: p.Gly585Val) and may also cause loss of expression in addition to reduced/absent activity (P: p.Lys429Asn, p.Ala772Pro, p.Tyr639Cys, p.Trp702Arg, p.Asp825His; N: p.Ala544Pro, p.Gly447Arg, p.Gly447Ala, p.Gly447Glu, p.Arg588Trp, p.Arg625Trp) (Choi et al., 2019; Guissart et al., 2020; Heidari et al., 2021; Matsell et al., 2024; Mohamadian et al., 2020). p.Lys429Asn and p.Lys429Met occur directly adjacent to the D428 active site (Palmgren & Nissen, 2011) (Choi et al., 2019). A variant in the A domain (p.Ile215Leu), however, did not disrupt ATP8A2 stability or activity (Guissart et al., 2020). The p.Leu538Pro variant identified in the present cases results in near total loss of ATP8A2 expression, likely due to the proline substitution breaking the alpha helical structure where this residue resides. Based on our findings and the findings of others, it is evident that the N and P domains are mutational hotspots, and missense variants in these domains often profoundly impact ATP8A2 stability and function. One recent study by Morgansen et al. simulated ATPase activity in 130 ATP8A2 missense variants in the M1-M4 transmembrane segments (Mogensen et al., 2024), which could have applicability for prioritizing missense variants within the catalytic domains for experimental validation in the future, as many variants of uncertain significance (VUS) remain unsolved.

Since many missense variants in the catalytic domains have been shown to result in loss of function, genotype-phenotype correlations in ATP8A2-related disease are complex. For example, of all of the missense variations in the cytoplasmic catalytic domains, normal brain MRI was only observed in a patient with the p.Trp702Arg variant while all others showed some degree of abnormality, including the two siblings reported in this case study (Guissart et al., 2020; Heidari et al., 2021; Martín-Hernández et al., 2016; McMillan et al., 2018; Mohamadian et al., 2020). This was in contrast to several reported frameshift and nonsense variants where brain MRI was normal in some or all patients (p.Arg581*, p.Asn596fs; p.Pro492_Ala554del; p.Arg588fs) (Alsahli et al., 2018; Alves Corazza et al., 2023; McMillan et al., 2018; Quintas, 2017), furthering highlighting the severity of ATP8A2-related disease that can stem from missense variants in the catalytic domains. There is also drastic variability in the brain manifestations that are reported across patients with *ATP8A2* variants, which correlates strongly to the broad expression pattern of ATP8A2 in many brain regions (Cacciagli et al., 2010; Onat et al., 2013). Interestingly, in the patients reported in this study, DWI revealed bilateral symmetric internal capsule hyperintensities that have not been previously reported in any patient with *ATP8A2* variants, which could explain documented sensorimotor deficits. These findings indicate restricted capsular water diffusion which can be caused by ischemia, inflammation, increased cell turnover, but most likely represent ongoing microstructural axonal dysfunction.

Compared to other cases in the literature, our patients exhibited many features shared by most other patients including developmental delay, intellectual disability, and dysarthria (Narishige et al., 2022). However, they presented with a milder clinical manifestation comparable to two families reported by Guissart et al. harboring missense variants in the N domain near the p.Leu538Pro variant. One family with two siblings carried the p.Gly585Val variant in homozygosity and presented with mild cerebellar ataxia. They were ambulant with ataxic gait. Mild cerebellar atrophy was observed on their MRI. They did not have encephalopathy, but had dysmetria, dysarthria, nystagmus, optic atrophy, tremors, and head titubation. The other family had one child with the p.Arg588Trp variant in homozygosity and presented with transient encephalopathy, developmental delay, hypotonia, nystagmus, dysmetria, and dysarthria, along with hearing loss. However, the child was able to stand and walk with aid (Guissart et al., 2020). Our patients presented with many of the same features, though neither one had optic atrophy or cerebellar atrophy, and P2 did not manifest ataxia.

The findings of this study and review of reported variation in ATP8A2 highlights how missense variation in the catalytic domains of this gene should not be overlooked in next generation sequencing studies. In fact, we believe that the combined knowledge of pathogenic phenotypes associated with catalytic domain missense changes could warrant reassessment of pathogenicity classification of many VUSs. Overall, the cases reported in this study introduce novel brain MRI findings into the clinical spectrum of ATP8A2-related disease, and further highlight the role of *ATP8A2* missense variants as a common mechanism.

## Author Contributions

KPF, SS, and MCM designed the study; AY, AAAH, LEOE, and MCM enrolled the research participants; MK, RA, FA, and AY collected the samples and relevant data; MAS, AY, VB, AAAH, INM, and MAE collected and reviewed the clinical information; KPF, SS, and MCM analyzed the sequencing data; EM and RSM performed and analyzed the protein expression experiments; KPF, SS, EM, RSM, and MCM wrote the manuscript; all authors reviewed the final manuscript.

## Acknowledgements

We thank the family for their participation in our study.

## Data Availability Statement

Data reported in this study will be made available upon request from the corresponding author.

